# Linking Trials to Publications: Enhancing Recall by Identifying Trial Registry Mentions in Full-Text

**DOI:** 10.1101/2025.06.09.25329285

**Authors:** Arthur W. Holt, Neil R. Smalheiser

## Abstract

We have developed a free, public web-based tool, Trials to Publications, https://arrowsmith.psych.uic.edu/cgi-bin/arrowsmith_uic/TrialPubLinking/trial_pub_link_start.cgi, which employs a machine learning model to predict which publications are likely to present clinical outcome results from a given registered trial in ClinicalTrials.gov. The tool has reasonably high precision, yet in a recent study we found that when registry mentions are not explicitly listed in metadata, textual clues (in title, abstract or other metadata) could identify only roughly 1/3–1/2 of the publications with high confidence. This finding has led us to expand the scope of the tool, to search for explicit mentions of registry numbers that are located within the full-text of publications. We have now retrieved ClinicalTrials.gov registry number mentions (NCT numbers) from the full-text of 3 online biomedical article collections (open access PubMed Central, EuroPMC, and OpenAlex), as well as retrieving biomedical citations that are mentioned within the ClinicalTrials.gov registry itself. These methods greatly increase the recall of identifying linked publications, and should assist those carrying out evidence syntheses as well as those studying the meta-science of clinical trials.

**Highlights:**

- Those conducting systematic reviews, other evidence syntheses, and meta-science analyses often need to examine published evidence arising from clinical trials. Finding publications linked to a given trial is a difficult manual process, but several automated tools have been developed. The Trials to Publications tool is the only free, public, currently maintained web-based tool that predicts publications linked to a given trial in ClinicalTrials.gov.
- A recent analysis indicated that the Trials to Publications tool has good precision but limited recall. In the present paper, we greatly enhanced the recall by identifying registry mentions in full-text of articles indexed in open access PubMed Central, EuroPMC and OpenAlex.
- The tool now has reasonably comprehensive coverage of registry mentions, both for identifying articles that present trial outcome results and for other types of articles that are linked to, or that discuss, the trials. This should greatly save effort during web searches of the literature.

## Introduction

One of the key steps in conducting a systematic review is to identify all of the trials and all of the corresponding publications relevant to a given clinical question. Although clinical trials may be registered in ClinicalTrials.gov and international registries, it is not an easy task to link a given trial to its publications. Only about half of trials give rise to any publications, and of those that do, only roughly half mention the trial registry number in the publication’s abstract or metadata [1-5]. Manual searching for linked publications is time-consuming and uncertain [6], leading several groups to devise machine learning-based models to predict which publications are likely to be linked to a given trial [7-12].

Our group has created several free, public web-based tools to assist evidence synthesis groups in identifying publications that arise from the same underlying trial [13, 14] and to predict which publications are likely to present clinical outcome results from a given registered trial [12]. The latter tool, called Trials to Publications [https://arrowsmith.psych.uic.edu/cgi-bin/arrowsmith_uic/TrialPubLinking/trial_pub_link_start.cgi], has reasonably high precision, yet in a recent study we found that when registry mentions are not listed in metadata, textual clues (in title, abstract or other metadata) could identify only roughly 1/3–1/2 of the publications with high confidence [15]. This finding has led us to expand the scope of the tool, to search for explicit mentions of registry numbers that are located within the full-text of publications.

Mentions that occur within the Methods section of articles are likely to present clinical trial outcome results [15], whereas mentions that occur in other sections tend to arise from review articles and diverse other article types.

Here we have extended the Trials to Publications tool by retrieving ClinicalTrials.gov registry number mentions (NCT numbers) from the full-text of 3 online biomedical article collections (PubMed Central (PMC), Europe PMC (EuroPMC), and OpenAlex), as well as retrieving citations that are mentioned within the ClinicalTrials.gov registry itself. These methods greatly increase the recall of identifying linked publications, and they are now displayed in the online Trials to Publications tool.

## Methods

### Predictive scores using the Trials to Publication model and tool [12]

Each registered trial in ClinicalTrials.gov, indicated with a primary completion date by the end of 2020, was compared to a candidate list of the 5,000 most similar PubMed articles as previously described, and a machine learning model was applied to predict the probability that the article reports clinical outcome results from that trial [12]. Results were stored in a relational database for look-up. User queries in the Trials to Publications tool, i.e. inputting a NCT number, also initiated real-time calculations to take into account any articles not pre-computed.

### Extracting registry mentions from PubMed articles

Metadata records of articles indexed in PubMed were downloaded in XML format. Registry mentions from ClinicalTrials.gov and 16 international registries were extracted from the abstract and other metadata fields using regular expressions that allow for a variety of common lexical variants in the way that NCT numbers are written [12], and stored in a relational database.

### Extracting registry mentions from open access PubMed Central articles

Articles indexed in PubMed Central (Open Access and Manuscript subsets) were downloaded in XML format as of January 2025. Article sections, including the Methods section, were identified using a custom program [15]. Registry mentions from ClinicalTrials.gov and 16 international registries were extracted from full-text using regular expressions that allow for a variety of common lexical variants [12].

### Extracting NCT mentions from all PubMed Central articles

We performed real-time queries via API to EuroPMC, a mirror site of PubMed Central that permits full-text search, anywhere in the article as well as restricted to the Methods section. Queries included the three most common lexical variants of NCT numbers, namely, NCTxxxxxxxx, NCT xxxxxxxx, and NCT:xxxxxxxx.

### Extracting NCT mentions from all articles indexed in OpenAlex

To identify NCT numbers mentioned in any portion of articles indexed in OpenAlex.org, we performed real-time full-text queries via API including the three most common lexical variants of NCT numbers. It was not possible to restrict search to the Methods section.

### Extracting biomedical article citations from ClinicalTrials.gov registries

Articles listed under Publications were extracted if their publication date was at least one year later than the official start date of the trial. We separately listed articles that were indicated as *Trial Results* vs. other articles that may have provided *Trial Background* information.

## Results

The tool is applicable for all trials in ClinicalTrials.gov and all publications indexed in PubMed, PubMed Central, and OpenAlex.org. However, for the results discussed here, we only consider a fixed corpus, namely, those 289,506 trials in ClinicalTrials.gov that were completed prior to January 2021, since more recent trials might not have had adequate time to obtain results and create publications. For this corpus, there are 125,154 articles in PubMed that explicitly mention one or more NCT numbers in metadata. An additional 25,526 articles are listed in the results publication field of the ClinicalTrials.gov record. The Trials to Publications machine learning-based model [12] identified 257,364 additional articles in PubMed that are predicted to be linked to a specific NCT number with high confidence (i.e., predicted probability >95.6%). Thus, prior to the present study, the Trials to Publications tool displayed or predicted a total of 408,044 articles.

To estimate the increase in recall produced by incorporating articles retrieved from open-access PMC, EuroPMC and OpenAlex.org searches, we randomly chose 20,000 registered trials in ClinicalTrials.gov (having a primary completion date between January 2002 and January 2021) for comparison of publications retrieved according to search source.

As shown in Figure 1 and Table 1, each online source showed some overlap with the others, but each contributed some unique articles. Overall, incorporating all “high precision” sources (i.e., Metadata, Trial Results, and the Methods section of open-access PMC and EuroPMC) increased recall by 45/37 = 21.6% relative to the previous baseline. In contrast, incorporating all sources increased recall by 62/37 = 67.6%, although many of the publications found with full text searches are reviews, ancillary publications, or other articles that discuss the research.

**Table 1.** Co-occurrence of Trial-Article Pairs by Search Source for 20,000 Trials. Each of the eight sources for searching for trial result publications identifies unique articles linked to each trial.

|  | Metadata | Trial Results | Model Predicted | Open PMC Methods | EuroPMC Methods | EuroPMC Full Text | Trial Background | OpenAlex Full Text |
| --- | --- | --- | --- | --- | --- | --- | --- | --- |
| **Metadata** | 100% | 5% | 16% | 22% | 25% | 80% | 2% | 43% |
| **Results in Trial Record** | 22% | 100% | 17% | 8% | 9% | 25% | 0% | 21% |
| **Model Predicted** | 8% | 2% | 100% | 3% | 4% | 9% | 1% | 7% |
| **Open PMC Methods** | 37% | 3% | 11% | 100% | 98% | 95% | 1% | 62% |
| **EuroPMC Methods** | 29% | 3% | 9% | 66% | 100% | 92% | 1% | 58% |
| **EuroPMC Full Text** | 20% | 1% | 5% | 14% | 20% | 100% | 1% | 46% |
| **Trial Background** | 7% | 0% | 4% | 3% | 4% | 9% | 100% | 7% |
| **OpenAlex Full Text** | 10% | 1% | 4% | 9% | 12% | 44% | 1% | 100% |

**Figure 1.**
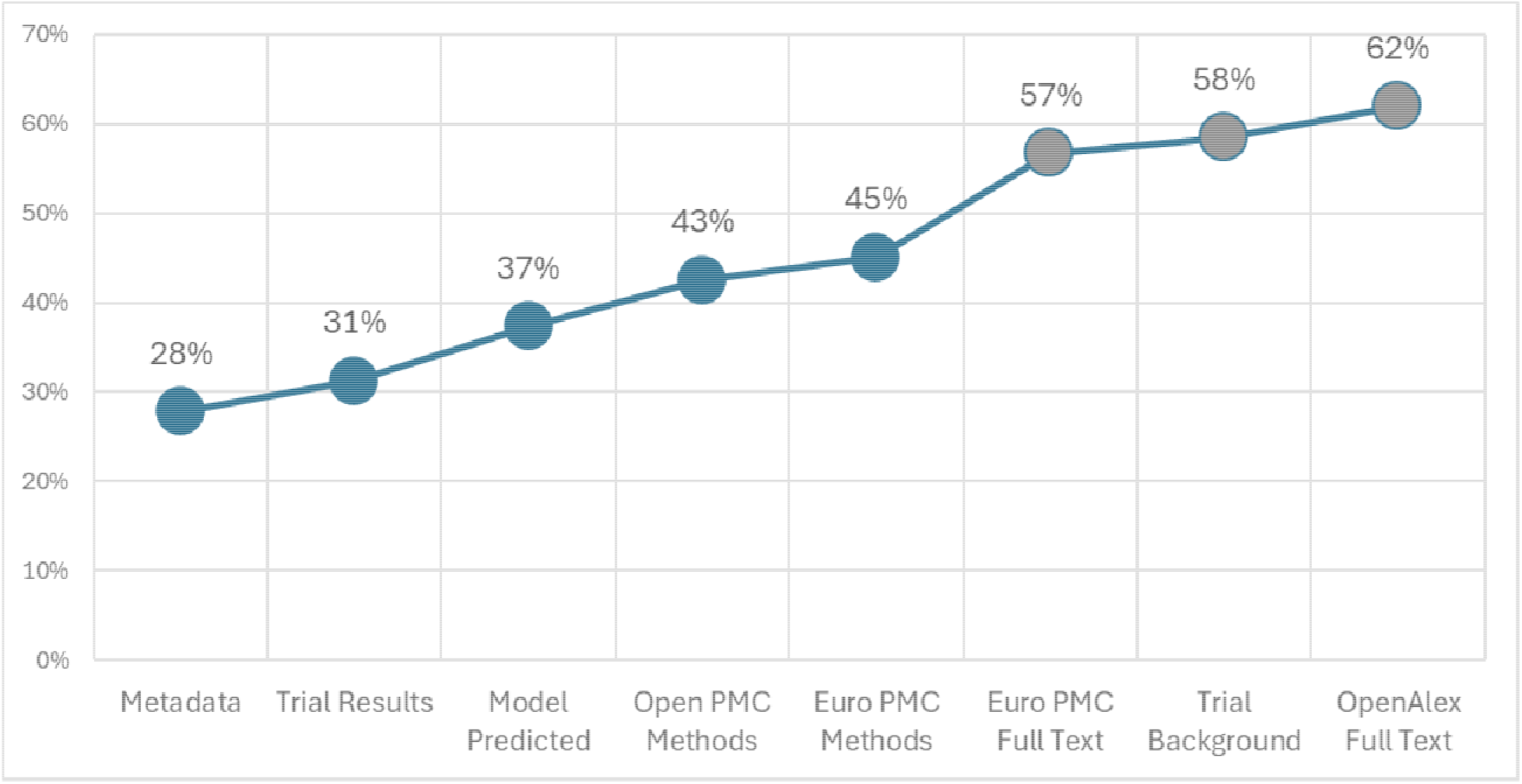
Cumulative % of Trials with at least One Registry Mention by Source

Interestingly, OpenAlex does not appear to have full coverage of PubMed or PubMed Central article texts, since only 43% and 46% of NCT mentions retrieved from PubMed and EuroPMC, respectively, were also retrieved from queries to OpenAlex.org with full text search.

## Discussion

When a user makes a query in the Trials to Publications tool by inputting a NCT number, the tool retrieves articles which are collated and displayed in two lists: The first consists of articles predicted to be linked to the registered trial according to the machine learning-based model, plus articles retrieved from the “high precision” sources, regarded as having relatively high precision for identifying linked articles that present clinical trial outcome results [12, 15]. The second list indicates articles retrieved from the additional “high recall” sources. The second list extends recall but is heterogeneous and contains some clinical trial articles together with e.g., review articles, book chapters, and preclinical studies. For articles lacking PMIDs, the publisher DOI is given.

How close does the expanded tool come to identifying ALL publications linked to registered trials in ClinicalTrials.gov? Based on our random sample of 20,000 trials, 9,008 or 45.04% had one or more publications identified from our high-precision sources, and of these, there were an average of 4 publications per trial (most trials had only one publication but there was a long tail of trials having many linked articles). This suggests that at least 9,008 × 4 = 36,032 articles are linked to the 20,000 trials, an under-estimate since not all linked articles mention any NCT numbers at all. On the other hand, 75,996 articles were retrieved from all 8 online sources, which is an over-estimate of the true number of linked articles since many of them are discussing trials rather than reporting trial outcome results. We estimate that at least 46% of articles having NCT mentions are truly linked to the registered trial, based on our previous data showing that high-precision mentions in (metadata+methods) comprised about 46% of all full-text mentions among open-access PMC articles [15]. Taking these numbers together, a back-of-the-envelope calculation suggests that estimated recall = (75,996 × 0.46) / 36,032 = 97.0%. This suggests that the great majority of linked articles are identified in the expanded tool. The remaining publications are likely to consist of articles that are published in grey literature or indexed in regional databases not covered by OpenAlex, as well as articles that are not identified by the machine-learning model that fail to mention a registry number at all.

No single method of searching the literature suffices to identify all linked publications. One may need to search grey literature [16], follow citation trails, or even write the authors of trials.

However, the expanded Trials to Publications tool provides rapid and reasonably comprehensive identification of articles linked to ClinicalTrials.gov registered trials that includes data across PubMed, PubMed Central, and OpenAlex.org. We hope that this will facilitate evidence syntheses as well as metascience studies of the clinical trial enterprise [17, 18].

## Author Contributions

AWH - Data curation; formal analysis; visualization; resources; writing - review & editing.

NRS - Conceptualization; methodology; writing - original draft; writing - review & editing; funding acquisition.

## Data Availability

Queries can be made to the free, public Trials to Publications tool without registration or login required https://arrowsmith.psych.uic.edu/cgi-bin/arrowsmith_uic/TrialPubLinking/trial_pub_link_start.cgi. Individual NCT numbers and batches of multiple numbers are both accepted.

## References

1. Manzoli L, Flacco ME, D’Addario M, Capasso L, De Vito C, Marzuillo C, Villari P, Ioannidis JP. Non-publication and delayed publication of randomized trials on vaccines: survey. BMJ. 2014 May;16(348): g3058. 10.1136/bmj.g3058.

2. Sreekrishnan A, Mampre D, Ormseth C, Miyares L, Leasure A, Ross JS, Sheth KN. Publication and dissemination of results in clinical trials of neurology. JAMA Neurol. 2018 Jul 1;75(7):890–1. 10.1001/jamaneurol.2018.0674.

3. Ross JS, Mulvey GK, Hines EM, Nissen SE, Krumholz HM. Trial publication after registration in ClinicalTrials.gov: a cross-sectional analysis. PLoS medicine. 2009;6(9):e1000144. 10.1371/journal.pmed.1000144

4. Huser V, Cimino JJ. Linking ClinicalTrials.gov and PubMed to track results of interventional human clinical trials. PloS one. 2013;8(7):e68409. 10.1371/journal.pone.0068409

5. Bashir R, Bourgeois FT, Dunn AG. A systematic review of the processes used to link clinical trial registrations to their published results. Syst Rev. 2017 Dec;6:1–7. 10.1186/s13643-017-0518-3.

6. Huser V, Cimino JJ. Precision and negative predictive value of links between ClinicalTrials.gov and PubMed. AMIA Annu Symp Proc. 2012;2012:400–8.

7. Goodwin TR, Skinner MA, Harabagiu SM. Automatically linking registered clinical trials to their published results with deep highway networks. AMIA summits on translational science proceedings. 2018;2018:54.

8. Dunn AG, Coiera E, Bourgeois FT. Unreported links between trial registrations and published articles were identified using document similarity measures in a cross-sectional analysis of ClinicalTrials.gov. J clin epidemiol. 2018:95:94–101. 10.1016/j.jclinepi.2017.12.007

9. Liu S, Bourgeois FT, Dunn AG. Identifying unreported links between ClinicalTrials.gov trial registrations and their published results. Res Synth Methods. 2022 May;13(3):342–352. doi: 10.1002/jrsm.1545.

10. Altman DG, Furberg CD, Grimshaw JM, Shanahan DR. Linked publications from a single trial: a thread of evidence. Trials. 2014 Dec;15:1–3. 10.1186/1745-6215-15-369.

11. Pan E, Roberts K. Linking cancer clinical trials to their result publications. AMIA summits on translational science proceedings. 2024;2024:642.

12. Smalheiser NR, Holt AW. A web-based tool for automatically linking clinical trials to their publications. J Am Med Inform Assoc. 2022 May 1;29(5):822–30. 10.1093/jamia/ocab290.

13. Shao W, Adams CE, Cohen AM, Davis JM, McDonagh MS, Thakurta S, Yu PS, Smalheiser NR. Aggregator: a machine learning approach to identifying MEDLINE articles that derive from the same underlying clinical trial. Methods. 2015 Mar;74:65–70. doi: 10.1016/j.ymeth.2014.11.006.

14. Smalheiser NR, Holt AW. New improved Aggregator: predicting which clinical trial articles derive from the same registered clinical trial. JAMIA Open. 2020 Oct 28;3(3):338–341. doi: 10.1093/jamiaopen/ooaa042.

15. Holt AM, Troy AM, Smalheiser NR. Distribution of trial registry numbers within full-text of PubMed Central articles: implications for linking trials to publications and indexing trial publication types. Trials. 2025 Jan 31;26(1):34. doi: 10.1186/s13063-025-08741-w.

16. Briscoe S, Nunns M, Shaw L. How do Cochrane authors conduct web searching to identify studies? Findings from a cross_Jsectional sample of Cochrane Reviews. Health Information & Libraries Journal. 2020 Dec;37(4):293–318.

17. Vorland CJ, Brown AW, Kilicoglu H, Ying X, Mayo-Wilson E. Publication of Results of Registered Trials With Published Study Protocols, 2011-2022. JAMA Netw Open. 2024 Jan 2;7(1):e2350688. doi: 10.1001/jamanetworkopen.2023.50688.

18. Lu J, Xu BB, Shen LL, Wu D, Xue Z, Zheng HL, Xie JW, Wang JB, Lin JX, Chen QY, Cao LL. Characteristics and research waste among randomized clinical trials in gastric cancer. JAMA Network Open. 2021Sep 1;4(9):e2124760.

